# Array Genotyping of Transfusion Relevant Blood Cell Antigens in 6946 Ancestrally Diverse Subjects

**DOI:** 10.1101/2025.04.25.25326432

**Authors:** Nicholas S. Gleadall, Lianne Koets, Olga Shamardina, Jeremy Gollub, Aaron J. Gottschalk, Orod Razeghi, Gorka Ochoa-Garay, Jonathan Stephens, Ram Varma, Jennifer Martin, Elias Allara, Colin J. Brown, James Daly, Emanuele Di Angelantonio, Shane Grimsley, W. Martin Howell, Kati Hyvärinen, Ute Jentsch, Nathalie Kingston, Celina Montemayor, Celeste Moya-Valera, John Ord, Jukka Partanen, David Roberts, Kathleen E. Stirrups, Sunitha Vege, Lindsay Walker, Andrea Harmer, Shantanu Kaushikkar, Willem H. Ouwehand, C. Ellen van der Schoot, Connie M. Westhoff, Blood transfusion Genomics Consortium, Barbera Veldhuisen, William J. Lane

**Author notes:** An additional list of collaborators who have contributed to the study as part of the ‘Blood transfusion Genomics Consortium’ is available in the supplemental Information. Joint first authors (Nicholas S. Gleadall, Lianne Koets). Joint last authors (Barbera Veldhuisen, William J. Lane).

## Abstract

**Key Points:** Array genotyping enables extensive blood cell antigen typing with >99.9% reproducibility across international laboratories.

Accurate antigen genotyping by array in diverse populations provides an opportunity to reduce alloimmunization by extended matching.

Category: Transfusion Medicine

Blood transfusions save millions of lives worldwide each year, yet formation of antibodies against non-self antigens remains a significant problem, particularly in frequently transfused patients. We designed and tested the Universal Blood Donor Typing (UBDT_PC1) array for automated high-throughput simultaneous typing of human erythroid, platelet, leukocyte, and neutrophil antigens (HEA, HPA, HLA, and HNA, respectively) to support selection of blood products matched beyond ABO/Rh. Typing samples from 6946 donors of European, African, Admixed American, South Asian, and East Asian ancestry at two different laboratories showed a genotype reproducibility of ≥99% for 17 244 variants, translating to 99.98%, 99.90%, and 99.93% concordance across 338 372 HEA, 53 270 HPA, and 107 094 HLA genotypes, respectively. Compared to previous clinical typing data, concordance was 99.9% and 99.6% for 245 874 HEA and 3726 HPA comparisons, respectively. HLA types were 99.1% concordant with clinical typing across 8130 comparisons, with imputation accuracy higher in Europeans versus non-Europeans. Seven variant *RHD* alleles, a *GYPB* deletion underlying the U− phenotype, and 14 high-frequency antigen negative types were also detected. Beyond blood typing, hereditary hemochromatosis-associated *HFE* variants were identified in 276 donors. We found that the UBDT_PC1 array can reliably type a wide range of blood cell antigens across diverse ancestries. Reproducibility and accuracy were retained when transfusion-relevant targets from the UBDT_PC1 array were incorporated into the UKBB_v2.2 genome-wide typing array. The results represent the potential for significant advancement towards improved patient care by reducing harm in transfused patients through extended matching.

## Introduction

Blood transfusion is a commonly administered therapy, serving as a life-saving intervention for millions of patients annually.^1^ To prevent acute hemolytic transfusion reactions (HTRs) and D immunization, patients are transfused with ABO- and RhD-compatible blood. However, alloimmunization to other blood group antigens still occurs in 2-5% of recipients.^2,3^ Patients undergoing chronic transfusions, i.e. those with sickle cell disorder, are at risk of alloimmunization, with rates ranging from 17% to 43%, depending on frequency of transfusion, use of extended matching, and local healthcare settings.^2–5^ In high-income countries, up to 10% of collected units are allocated to patients requiring frequent transfusions, including those with inherited anemias like sickle cell disorder and thalassemia.^6^ For these patients, guidance stipulates that blood for transfusion should be matched for the C/c, E/e, and K antigens to reduce the risk of alloimmunization, however, this is not always applied consistently, and blood is not routinely matched for other human erythroid antigens (HEA), which can also elicit antibody formation.^7–10^

Alloimmunization, as a consequence of limited antigen matching in transfusion, leads to increased risk of HTRs, delays in securing compatible blood, challenges in maintaining a chronic transfusion program, increased costs, and an increased risk of hyperhemolysis, disproportionately affecting those with sickle cell disorder.^11–14^ Similarly, frequent platelet transfusions can cause alloantibody formation, generally against human leukocyte antigens (HLA) and, in some cases, human platelet antigens (HPA), rendering standard platelet selection ineffective.^15,16^ Furthermore, antibodies against human neutrophil antigens (HNA) may cause transfusion-related acute lung injury, and transfusing blood mismatched for HLA may lead to increased rejection in kidney transplant recipients.^17–19^

Comprehensive extended matching between patients and donors significantly reduces the risk of developing antibodies against non-self antigens.^20^ While serologic typing has been the gold standard for HEA, DNA-based methods have been preferred for HPA, HNA and HLA typing for decades. Serologic extended typing is not amenable to high-throughput applications and typing reagents for several clinically relevant HEA types are either unavailable, scarce, or unreliable.^21–24^

The genetic basis of 366 HEA types, classified across 47 blood group systems^25^, is known and DNA-based tests using various genotyping techniques are commercially available for a limited number of HEA types.^26^ Currently, there is no single platform that can provide comprehensive typing of HEA, HPA, HNA, and HLA, creating an opportunity for technological innovation in transfusion medicine. Extended genotyping of blood group antigens has been shown to significantly reduce alloimmunization rates and improve transfusion outcomes, particularly for chronically transfused patients.^20^ Thus, the Blood transfusion Genomics Consortium (BGC), an international collaboration between 18 institutions (supplemental Table 1), was formed to develop a high-throughput genotyping array for comprehensive blood cell typing. The array design also includes blood product quality-related markers that impact donor health, component functionality, and overall transfusion safety.^27–32^

In this study, the BGC significantly expanded the genetic targets relevant for transfusion and increased the number and diversity of sample cohorts tested, building on a prior feasibility study using the Axiom^TM^ UK Biobank (UKBB) array.^33^ That study prompted further design and targeted enhancements to develop an array tailored to the needs of blood services called the Axiom^TM^ Universal Blood Donor Typing (UBDT_PC1) array. In this international multicentre study, we demonstrate that the UBDT_PC1 and UKBB_v2.2 arrays can be used for comprehensive typing in accredited laboratories with high accuracy and reproducibility when testing subjects of different ancestries.

## Methods

### Study subjects

DNA samples and associated metadata from 7279 study subjects were obtained from 7 blood services in compliance with the appropriate regulatory approvals (supplemental Information). Of these samples, 6946 were from blood donors explicitly collected for the study and preferentially selected due to having extended typing data available (Figure 1A). An additional 333 samples, with complex or rare blood cell antigen types, were retrieved from DNA repositories at NHSBT, Sanquin, and NYBC to ensure the inclusion of at least 5 homozygous samples per antigen type for validation purposes (supplemental Table 2A–C, supplemental Information).

**Figure 1:**
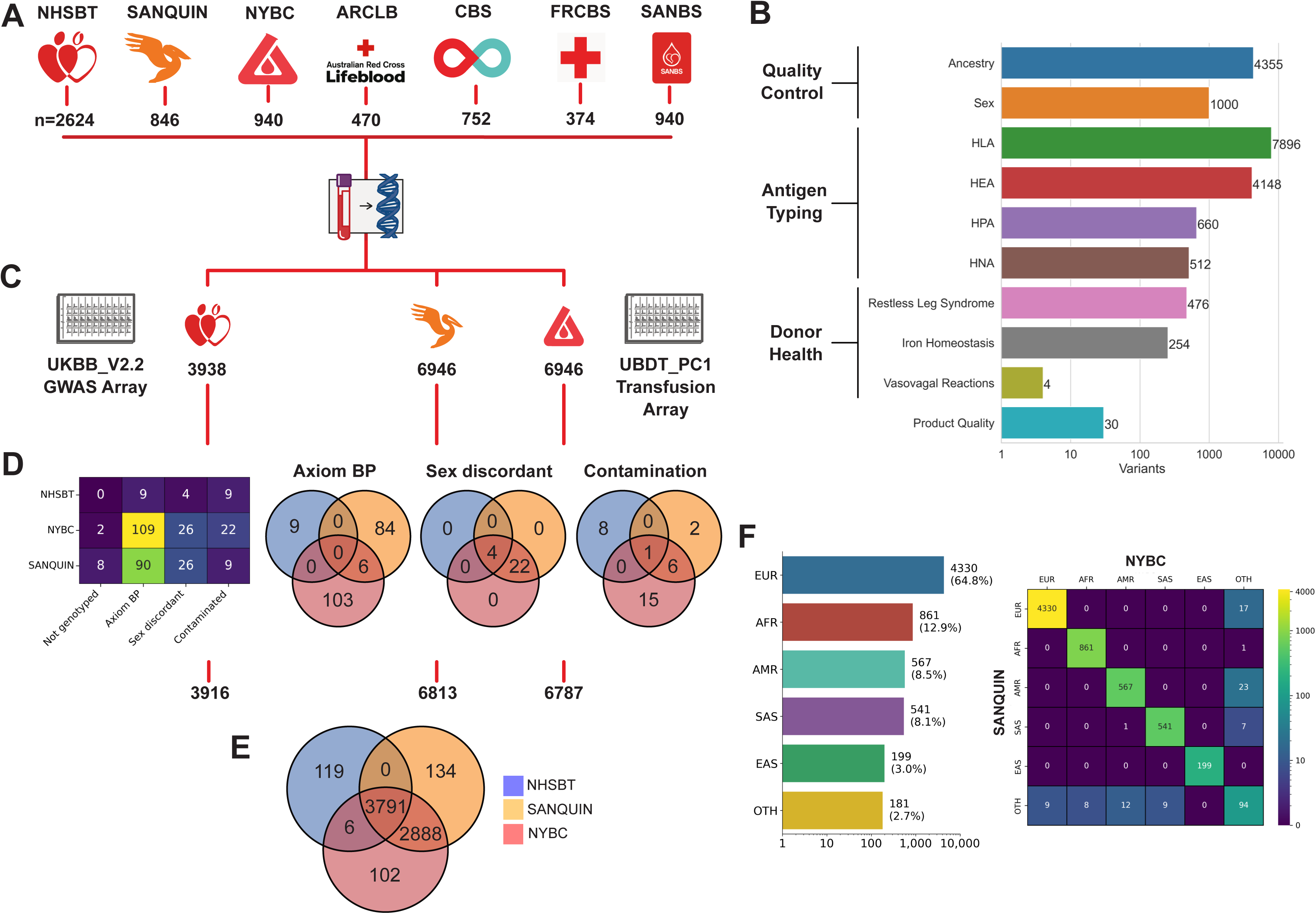
Study design. (**A**) Samples: Number of DNA samples provided by the 7 blood services (NHS Blood and Transplant (NHSBT); Sanquin Blood Supply Foundation (SANQUIN); New York Blood Center (NYBC); Australian Red Cross Lifeblood, (ARCLB); Canadian Blood Services (CBS); Finnish Red Cross Blood Service (FRCBS); South African National Blood Service (SANBS)). (**B**) Array content: Bar plot indicating the number of probes per category in the transfusion module. Human Leukocyte Antigens (HLA), Human Erythrocyte Antigens (HEA), Human Platelet Antigens (HPA), and Human Neutrophil Antigens (HNA). (**C**) Genotyping: 6946 identical DNA samples were genotyped with the UBDT_PC1 Transfusion Array at Sanquin and NYBC, with 3938 of these samples also genotyped using the UKBB_v2.2 GWAS Array by NHSBT. (**D**) Quality control (QC): Heatmap gives the reason for and number of samples failing QC for the 3 genotyping laboratories. Venn diagrams show overlap in samples that failed Axiom Best Practices (Axiom BP) QC, gender-vs-sex discordance (Sex discordant), and evidence of contamination (Contamination). (**E**) Venn diagram showing the overlap in samples passing QC. (**F**) Ancestry: (left) Barplot showing genetically inferred ancestry of samples typed successfully by Sanquin and NYBC (6679 samples). European (EUR), African (AFR), Admixed American (AMR), South Asian (SAS), East Asian (EAS) and Other (OTH) are shown. (right) Heatmap showing concordance between the ancestry inferred from the Sanquin and NYBC genotyping results, respectively.

### Axiom^TM^ array test

The sample preparation and hybridization protocols for the Axiom^TM^ array test were followed as previously described (supplemental Information).^34^ The UBDT_PC1 array contains 20 681 probes for genotyping 19 335 unique DNA variants, which are defined hereafter as the ‘transfusion module’ and are relevant to quality control (QC) (inference of ancestry and sex), antigen typing, donor health, and product quality (Figure 1B). The transfusion module has been modified since initial description to increase the number of HEAs, which can be typed from 22 to 51, with corresponding enhancements made to the bloodTyper application.^35^ The UKBB_v2.2 array includes 819 899 probes for typing 812 583 variants dispersed across the genome, including the transfusion module and variants required for genome-wide association studies (GWAS).^34^ An integrated analysis package (IAP) was developed, which applies multiple automated sequential algorithms for genotype QC followed by inference of HEA/HPA and HLA class I and II types using the bloodTyper and the HLA*IMP:02 model (Applied Biosystems HLA Analysis software v2.12.0RC1, supplemental Information).^35,37–39^

### Comparison of typing data

All 6946 study samples underwent genotyping in duplicate, performed once each by Sanquin and NYBC using the UBDT_PC1 array, while NHSBT conducted additional genotyping on a subset of 3938 samples with consent for genome-wide typing using the UKBB_v2.2 array (Figure 1C). The reproducibility of genotyping results between laboratories was assessed using in-house developed software. HEA, HPA, and HLA typing results retrieved from electronic donor records, hereafter referred to as clinical typing data (supplemental Table 3A), were compared with array-generated typing data. Sanquin and NYBC investigated discordances between HEA and HPA typing results, whereas the discordances in HLA types were investigated by NHSBT, using analysis of specific probe call plots and currently used accredited molecular assays (supplemental Table 3A).^40–44^

### Data availability

Requests for access to study data should be e-mailed to.

## Results

### Genotyping

Identical sets of 6946 DNA samples were distributed to Sanquin and NYBC, with a subset of 3938 samples also distributed to NHSBT (Figure 1A,C). 17 820 (99.94%) of the distributed samples were genotyped successfully, and 208 (1.17%) failed Axiom^TM^ Best Practice QC during genotype calling. Another 56 (0.31%) and 40 (0.22%) samples were excluded for gender vs. sex discordance or contamination, respectively (Figure 1D). Unified datasets were created using samples passing QC, one composed of 6679 samples typed in duplicate with the UBDT_PC1 array and another consisting of 3791 samples typed once using the UKBB_v2.2 array (Figure 1E).

### Ancestry inference

Genetic ancestry was inferred for each sample using principal component analysis and Gaussian Mixture Models fit on genotypes from the 1000 Genomes dataset.^36^ Ancestry results from the Sanquin and NYBC laboratories were concordant for 6594 (98.7%) of the 6679 samples. 84 of the discordant results were due to low-confidence (*P*<0.9) ancestry calls, resulting in ‘Other’ being assigned, and 1 was classified as South Asian by Sanquin versus Admixed American by NYBC, respectively (Figure 1F). 35.2% (n=2349) of the samples were from non-European subjects, with 21.4% (1428) being of African or Admixed American ancestry. Similarly, concordance for the 3791 DNA samples typed by NHSBT was 95.59% (167/3791), with 59 being discordant between the African and Admixed American populations and 108 being assigned as ‘Other’. 3770279

### Genotype reproducibility

Genotype reproducibility between Sanquin and NYBC exceeded 99% for 17 244 (83.4%) of 20 681 total probes in the unified dataset (n=6679) (Figure 2A). Statistical analysis revealed 14 368 (83.4%) of the 17 244 high-concordance probes showed no significant difference (Pearson test, P≥0.001) between minor allele frequency (MAF) in this study and those from the gnomAD v4.1.0 dataset, which were derived from whole genome sequencing results of 36 667 Europeans (Figure 2B).45 Among 59 antigen typing probes (51 HEA and 7 HPA probes), the genotypes of 50 were 100% reproducible, 7 showed ≥99%, and 2 variants (M/N and HPA-3) had <90% reproducibility (Figure 2C). HPA-3 results were therefore excluded from analysis, but M/N antigen typing results were retained despite <99% reproducibility for 1 of 3 probes as the bloodTyper algorithm considers multiple probes for interpretation. The transfusion module also includes 11 variants for direct HNA typing and 35 variants for ferritin polygenic score calculation (supplemental Table 7), with >99% reproducibility in 44 variants (Figure 2C).^32^

**Figure 2:**
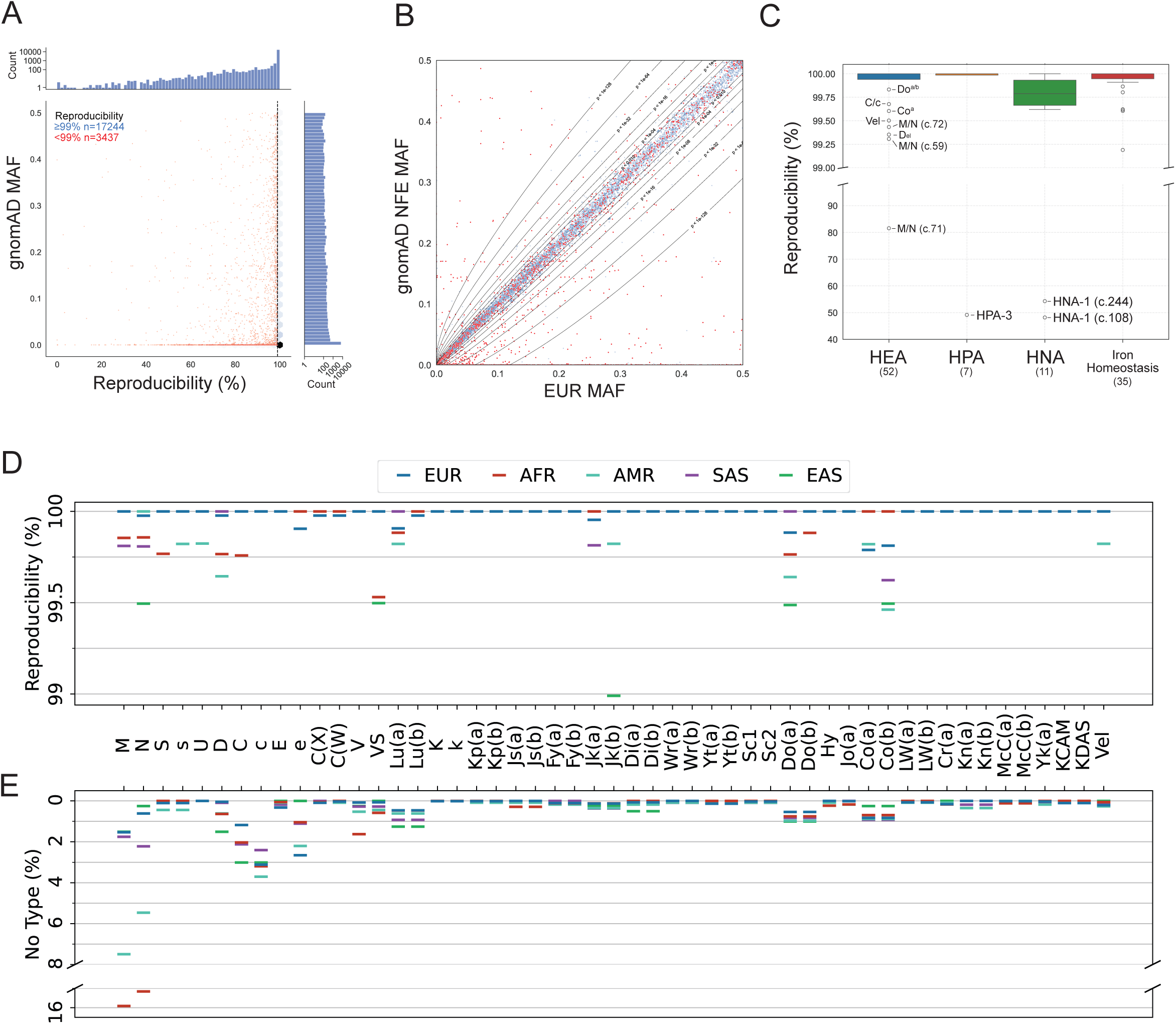
Reproducibility of typing results between Sanquin and NYBC for the 6679 DNA samples of the unified data. (**A**) Genotype reproducibility for 20 681 bi-allelic probe-variant pairs included in the UBDT_PC1 array design. Reproducibility expressed as percentage of concordant genotype comparisons, and gnomAD minor allele frequency (MAF) for each variant are displayed on the x- and y-axes, respectively. Blue hexagons and red dots on the central scatterplot represent the density of probes with reproducibility ≥99% and individual probes with <99% concordance, respectively. Marginal histograms show probe counts on a log scale. (**B**) Correlation of the MAF in EUR study subjects versus (non-Finnish) EUR subjects from the gnomAD database for each probe-variant pair. Probes with ≥99% and <99% genotype reproducibility are shown in blue and red, respectively. Contour lines represent boundaries of statistical significance with corresponding *P* values calculated using the X-square test. (**C**) Genotype reproducibility for critical blood antigen types and iron homeostasis probes. Box plots show the percentage reproducibility between genotypes, split across 2 y-axes ranges to highlight high-reproducibility results (99-100%) and broader distribution patterns (40-99%). Data are shown for Human Erythrocyte Antigens (HEA), Human Platelet Antigens (HPA), Human Neutrophil Antigens (HNA), and iron homeostasis variants in blue, orange, green, and red, respectively. Box plots display the median (centre line), interquartile range (IQR; box), whiskers (1.5×IQR), and outliers (black circles). Outlier variants are annotated with relevant antigen types. (**D**) Reproducibility between HEA types generated by the Sanquin and NYBC laboratories. The reproducibility is given as a percentage between on the y-axis for the 51 HEA types on the x-axis. Results are stratified for the 5 ancestry groups. Where the bars for different ancestries are at identical values, only one bar is shown in the order of the legend, i.e. blue for EUR subjects in most cases. (**E**) The percentage of no-type results is given on the y-axis for the 51 HEA types on the x-axis. HEA types with identical percentage of no-type results are visualised according to the principles of Figure 2D.

### HEA genotyping

Antigen genotype reproducibility between Sanquin and NYBC was 99.98% in 338 372 comparisons for 51 HEA across 6679 samples (Figure 2D). 31 HEA showed 100% type reproducibility, and 14 exceeded 99.75%. Types for N, VS, Jk^b^, Do^a^ and Co^b^ had ≤99.5% reproducibility. Significant (*P*≤0.05, Fisher’s Exact Test) reproducibility differences between Europeans and other ancestries were limited to: C and S antigens (African), D antigen (Admixed American), Jk^b^ antigen (East Asian), and VS antigen (African and East Asian). 2257 comparisons couldn’t be performed due to missing data in both replicates (n=559, 0.16%) or a single replicate (n=1698, 0.50%), primarily affecting African (n=224 [0.51%], n=345 [0.78%] of 43 911) and Admixed American samples (n=81 [0.28%], n=209 [0.72%] of 28 917). Ancestry-stratified typing density averaged 99.6% (99.2%-99.8%) for array typing versus 35.8% (29.4%-66.8%) for clinical typing (Figure 3A, supplemental Table 3B,C). Genotyping increased the number of available HEA types 2.72 times from 124 364 to 339 221 (supplemental Table 3B,C). All HEA types had missingness (no-type results) ≤2.75% except M, N, C, c, with M/N no-type results higher in Admixed American (7.5%, 5.5%) and African samples (15.9%, 15.1%) versus Europeans (1.53%, 0.61%) (Figure 2E).

**Figure 3:**
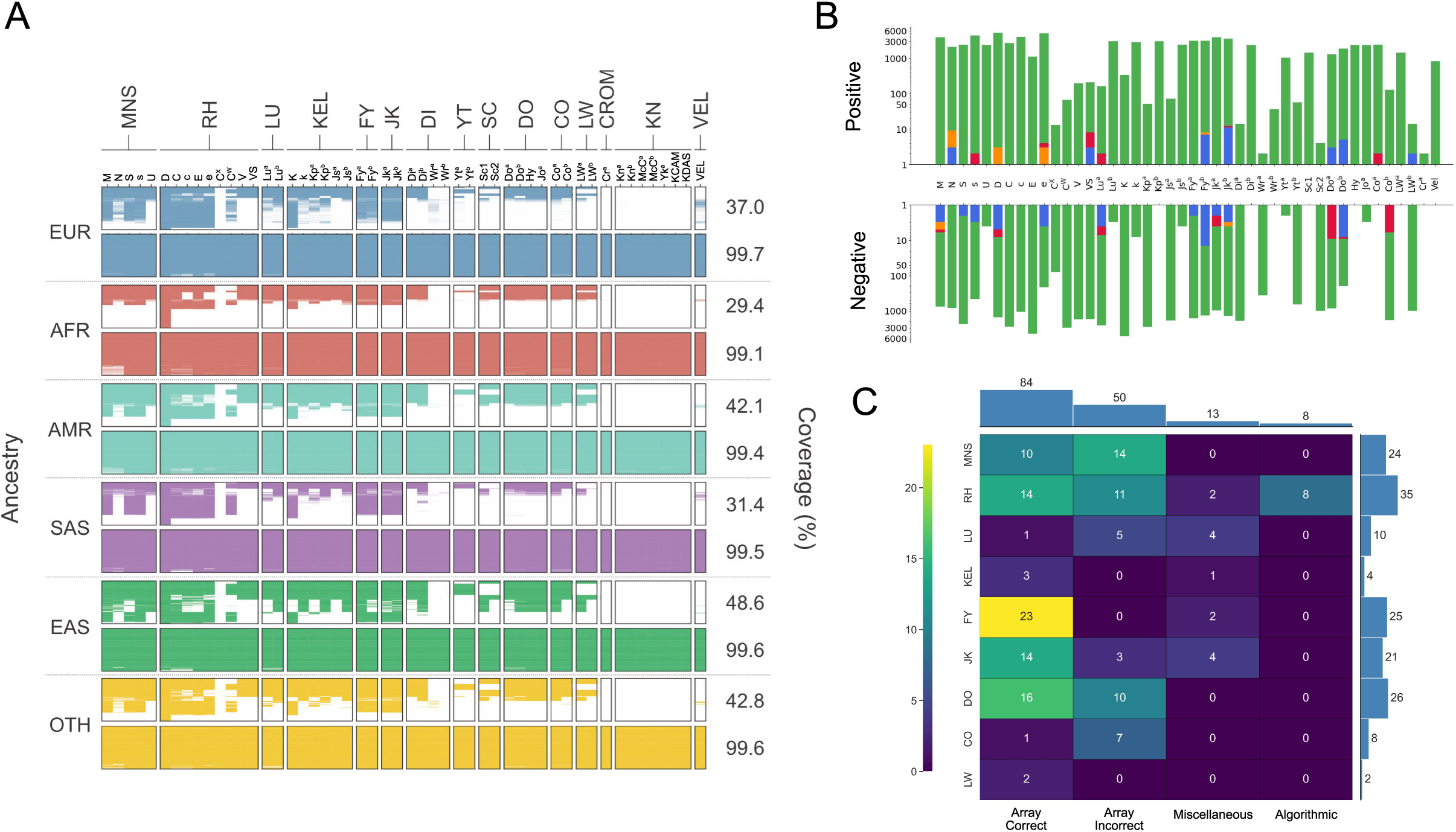
Array generated typing for Human Erythroid Antigens. (**A**) Comparison of clinical and UBDT_PC1 array-based Human Erythroid Antigen (HEA) typing density. In the graph, the presence of colour represents a typing result (positive or negative), and the absence of colour indicates a lack of a typing result. The graph is stratified according to the ancestry of the study subjects, with a top and bottom panel for each ancestry group representing the density of clinical and array tying results, respectively. European (EUR), African (AFR), Admixed American (AMR), South Asian (SAS), East Asian (EAS), and Other (OTH) ancestries are shown. The HEA systems and relevant antigen types are indicated on top of the graph, and the typing density as a percentage of the total possible types is given on the right of the graph. (B) Concordance between clinical and array-generated HEA types. Bar plots showing the number of comparisons (y-axis) per HEA type (x-axis) with concordant results obtained by both Sanquin and NYBC in green and discordant results by both Sanquin and NYBC in blue, Sanquin only in orange and NYBC only in red. Ascending and descending bars represent the number of comparisons to positive clinical or negative clinical antigen types, respectively. Bar plots show the number of comparisons on a log scale. (**C**) HEA typing discordances. Heatmap showing the cause of the discordance (columns) between clinical and array-generated types by HEA system (rows). The number of unique discordances per system and the number per cause of discordance are given on the right and top marginal bar plots, respectively.

### Concordance between clinical and array-derived HEA types

Concordance between clinical and array-generated typing results for the 6679 samples was calculated for HEA types generated using the UBDT_PC1 data from Sanquin and NYBC. Analysis was performed for 44 of the 51 antigens typed by the array as no clinical typing data was available for 7 KN system antigens (Kn^a^, Kn^b^, McC^a^, McC^b^, Yk^a^, KCAM, KDAS). Clinical and array-determined HEA types showed concordance rates of 99.91% (123 520 comparisons) tested at Sanquin and 99.89% (123 354 comparisons) at NYBC (Figure 3B, supplemental Table 4A,B). In the 246 874 total comparisons across both laboratories, 255 discordances (0.10%) were identified. These represented 155 unique sample-to-HEA-type discordances from 143 subjects; 11 DNA samples showed multiple discordances, primarily within the RH system. Of the total discordances, 100 were observed at both laboratories, while 16 were unique to Sanquin and 39 were unique to NYBC (Figure 3B, supplemental Table 4B). Investigation of discordances revealed 84 (54.2%) supported array-generated results, 58 (37.4%) were attributed to inadequate array or bloodTyper performance. Of the remaining 13 (8.4%) miscellaneous discordances, 11 were resolved: 7 were due to high-impact variants present on the array but not used by bloodTyper, 2 cases were likely due to incorrect clinical genotyping (pending confirmation), 1 was attributed to a novel variant absent from the array, and 1 was due to incorrect clinical data submission. Only 2 cases remained unresolved due to the lack of sufficient DNA for investigation (Figure 3C, supplemental Table 4B).

### Common, rare and complex HEA types

Array typing results confirmed expected blood group frequency variations between ancestral groups (Figure 4A). Briefly, the U− phenotype, resulting from homozygous *GYPB* deletion, occurred in 0.5% of African subjects (4/861).^46–48^ Dce haplotype homozygosity exhibited marked ancestry-dependent distribution: 40.8% in Africans, 3.5% in Admixed Americans, and 0.1% in Europeans. We observed significant ancestral differences in the *ACKR1* promoter variant (NM_002036.4:c.−67T>C) associated with the Fy(a−b−) phenotype (FY*01N.01 and FY*02N.01). 69.2% of African subjects (555/861) and 6.2% of Admixed Americans (35/567) carried the variant in homozygosity, while it was absent in European and Asian subjects. All 590 homozygous carriers were also homozygous for the NM_002036.4:c.125G>A (FY*02) variant located 671 base pairs downstream.^25,49^

**Figure 4:**
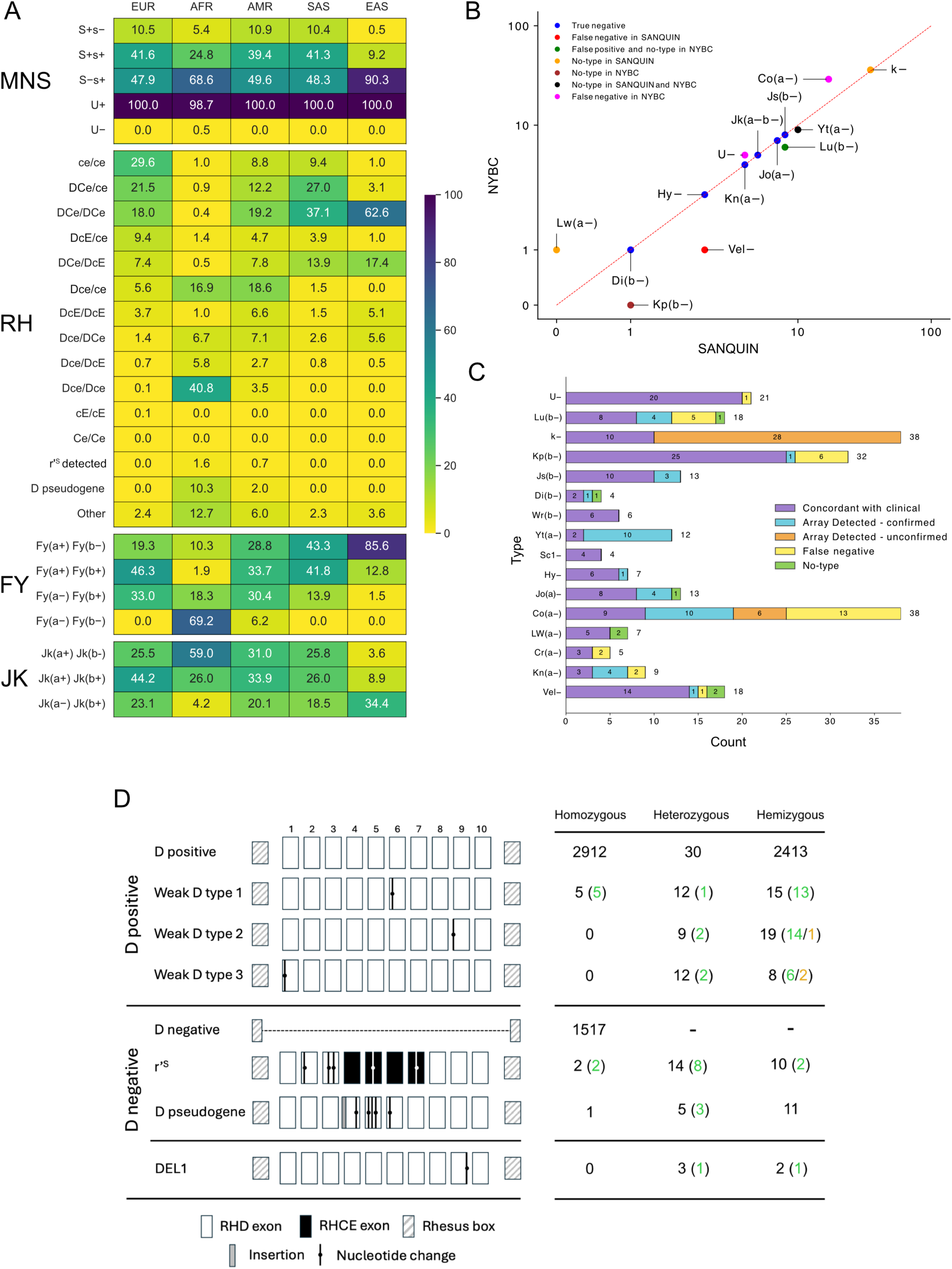
Common and rare Human Erythroid Antigen types. (**A**) Ancestral differences in frequencies of some common Human Erythroid Antigen (HEA) types which frequently elicit alloantibody formation. Heatmap with the ancestry stratified frequencies of the common MNS, RH, FY and JK types in the unified set of 6679 DNA samples. Heatmap colours range from yellow (0%) to deep blue (100%), showing HEA-type frequencies within each ancestry group. (**B**) Number of high-frequency antigen (HFA)-negative samples identified in the unified set of 6679 DNA samples with those identified by Sanquin and NYBC on the x- and y-axes, respectively. True negative, false negative in Sanquin, false positive and no-type in NYBC, no-type in Sanquin, no-type in NYBC, no-type in Sanquin and NYBC, and false negative in NYBC are showing in blue, red, green, orange, brown, black, and magenta, respectively. (**C**) Number of subjects typed negative for 16 HFA identified in the extended unified sample set. Bar plot shows phenotype and the count of negative typing results on the x- and y-axes, respectively. Typing results concordant with clinical type, array detected and confirmed, array detected and unconfirmed, false negative array types, and no-type results are shown in purple, blue, orange, yellow and green, respectively. (**D**) Concordance between clinical and array-generated results for DNA samples harbouring complex Rh genotypes. A graphical representation of 8 alleles of the *RHD* gene, in descending order: D positive (*RHD*01*), Weak D type 1 (*RHD*01W.1*), Weak D type 2 (*RHD*01W.2*), Weak D type 3 (*RHD*01W.3*), D negative (*RHD*01N.01)*, r’^S^ type 1 (*RHD*03N.01*), D pseudogene (*RHD*08N.01*), DEL1 (*RHD*01EL.01*). Counts on the right show the number of alleles detected, confirmed by clinical type, and discordant in the extended unified sample set in black, green, and orange, respectively.

The 6679 study samples contained a limited number of samples known to lack high-frequency antigen (HFA) negative phenotypes: U−, Lu(b−), k−, Kp(b−), Js(b−), Di(b−), Wr(b−), Yt(a−), Sc1−, Hy−, Jo(a−), Co(a−), LW(a−), Cr(a−), Kn(a−), and Vel−. Two approaches were taken to address this. First, 333 additional samples with rare HEA/HPA types determined by clinical typing were also genotyped by array. Of these, 131 were known to be HFA-negative (supplemental Table 2C). Second, array-derived genotypes among the 6679 samples identified 114 potential HFA-negative types, of which 90 (78.9%) were typed negative by both Sanquin and NYBC, 15 (13.2%) were typed HFA-negative by only 1 laboratory, and 9 (7.89%) did not produce a HEA type call in one replicate (Figure 4B). Of the 245 potential HFA-negative samples identified, 208 (84.9%) were concordant with previous clinical typing or confirmatory Sanger sequencing, 30 (12.2%) were discordant with clinical type or Sanger sequencing, and 7 (2.85%) failed to produce a HEA type by array due to a no-type result. Investigation of the 30 discordances revealed that 13 were correctly typed by array and 17 were typed false-negative by the array or bloodTyper algorithm; 12/17 as Co(a−) at the NYBC site due to failure of automated genotype calling (Figure 4C, supplemental Table 4C). In total, we observed 16 uncommon or rare HFA-negative types in the array data, resulting in the identification of 73 new HFA-negative donors.

The 333 additional samples also contained *RHD* alleles encoding common weak D, D negative, and D_el_ phenotypes previously identified through clinical genotyping and serology (Figure 4D). Of 63 known altered alleles, 60 (95.2%) were correctly genotyped (Figure 4D, supplemental Table 4D). Three - 1 weak D type 2 and 2 weak D type 3 - samples were missed due to failure of genotype calling, with 1 failing genotyping altogether and 2 samples having no-type results for the D antigen specifically. Subsequent repeat array testing yielded correct results for these samples (Figure 4D, supplemental table 4C).

Finally, 134 of the 333 additional known samples were selected either to increase the number of samples with clinical typing (n=90, e.g. Kn(a+)) or for which complex genetics underpinned HEA type expression (n=44, e.g. U+^var^). Concordance between array and clinical types was 93.3% (n=125), and of the remaining 9 samples, investigation supported the array result for 4; incorrect array call for 2, and no-type result for 3 (supplemental Table 4C).

### HPA typing

The reproducibility of array-inferred HPA antigen types was 99.88% in 53 270 comparisons of the 6679 samples. Ancestry-stratified frequencies of HPA-1,-2,-5,-15 align with expected frequencies (supplemental Table 4E).^50^ For the 8 HPA types, 99.58% concordance was observed across 3726 comparisons between clinical and array-generated types (supplemental Figure 1, supplemental Table 4D). At Sanquin, 6 (0.42%) discordant results were observed, with all but 1 of these also being observed by NYBC (supplemental Figure 1, supplemental Table 4D). Investigation of discordances revealed 4 (66.6%) supported array-generated results, 1 (16.7%) was attributed to inadequate array performance, and 1 (16.7%) remained unresolved. Finally, 10 of the 333 additional samples were selected for having rare HPA-1b,b, - 2b,b, or −5b,b types, and 100% concordance was observed (supplemental Table 4C).

In summary, analysis of the entire dataset enabled robust array validation for typing 44 HEA and 8 HPA where a minimum of 5 samples representing both negative and positive phenotypes for each possible type were available. Limited validation was achieved for several rare types, Js(b−), Di(b−), Sc1−, Cr(a−), and Kn(b−), with at least 1 clinically typed sample available for each. However, the limited availability of clinical typing prevented the validation of McC(a+) and Yk(a+) types (supplemental Table 4F).

### HLA

Many blood services type platelet donors for HLA class I and II antigens to provide HLA-matched platelets for refractory patients and to identify potential bone marrow registry volunteers. The transfusion module contains probes for 7896 variants across 6 Mb of the extended HLA region for typing (Figure 5A). Interlaboratory genotype reproducibility between Sanquin and NYBC for the 6679 unified samples was ≥99% for 6829 (86.5%) of the 7896 HLA locus variants (Figure 5B). HLA*IMP:02 was used to impute two-field class I and II types. Confidence score distributions between European and non-European subjects showed significant differences for some loci, with HLA-DPB1 having the lowest difference (Kolmogorov-Smirnov test statistic D=0.031, P=0.046) and HLA-B the largest (Kolmogorov-Smirnov test statistic D=0.154, P=6.51×10⁻³) (Figure 5C, supplemental Figure 2A). We observed 164 class I alleles (50 HLA-A, 84 HLA-B, 30 HLA-C) and 103 class II alleles (51 HLA-DRB1, 26 HLA-DQB1, 26 HLA-DPB1), with the 4 most frequent alleles per ancestry group shown in Figure 5C and supplemental Figure 2B.

**Figure 5:**
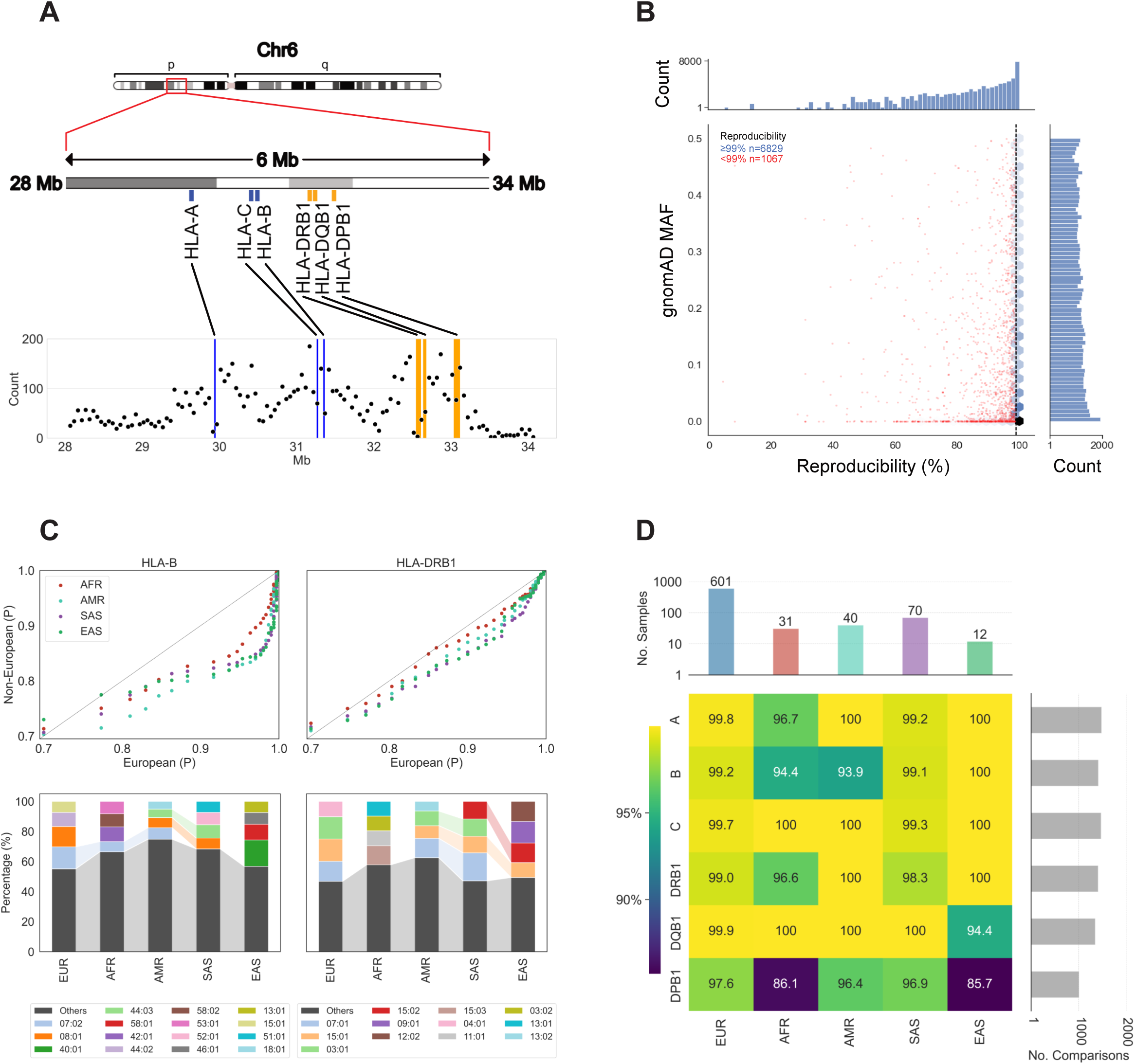
Human Leukocyte Antigen typing. (**A**) Schematic depicting density and location of probes across the Human Leukocyte Antigen (HLA) region, with upper panel: location of the extended HLA locus on chromosome 6p indicated by a red box window; middle panel: zoomed in visual of the 6 Mb extended HLA locus spanning from genome coordinates 28-34 Mb and depicting 2 sets of 3, class I and class II, genes, respectively; lower panel: graph visualising the number of probes per 50 000 base pair windows across the 6 Mb, with number of probes/window on the y-axis and genomic coordinates in Mb on the x-axis. (**B**) Genotype reproducibility for 7896 bi-allelic probe-variant pairs included in the UBDT_PC1 array design. Reproducibility expressed as percentage of concordant genotype comparisons, and gnomAD minor allele frequency (MAF) for each variant are displayed on the x- and y-axes, respectively. Blue hexagons and red dots on the central scatterplot represent the density of probes with reproducibility ≥99% and individual probes with <99% concordance, respectively. Marginal histograms show probe counts on a log scale. (B) Allele diversity: upper panel: Quantile-Quantile (Q-Q) plots illustrating the distribution of probability scores in calling alleles for European and non-European ancestries across the HLA-B (left) and HLA-DRB1 (right) genes. Quantiles of probabilities for European (EUR) and non-European samples [African (AFR), Admixed American (AMR), South Asian (SAS), and East Asian (EAS)] are shown on the x- and y-axes, respectively. Lower panel: Stacked bar charts showing the frequency distribution of the top 4 alleles for the different ancestry groups for the HLA-B (left) and HLA-DRB1 (right) genes, respectively. For both genes, the frequencies of the top alleles are normalised to percentage values (y-axis) and ancestry groups are given on the x-axis. Shaded lines are drawn between bar segments representing the same allele. (**D**) Concordance between clinical and HLA*IMP:02 imputed types are presented in a heatmap, expressed as an agreeing percentage of total comparisons ranging from dark blue (85.7%) to bright yellow (100%). The vertical and horizontal marginal bar plots give on the y-axis the number of samples used for the concordance analysis stratified per ancestry group on a log scale and the number of comparisons made for each of the 6 HLA genes.

Array-generated HLA type reproducibility was 99.93%, with only 70 discordances (0.07%) in 107 094 comparisons (supplemental Table 5A). For 766 of the 6679 (11.46%) subjects in the unified set clinical HLA typing results were available, including 164 (21.4%) non-European subjects (supplemental Tables 5B,C). Clinical and array-imputed HLA types showed 99.1% concordance, with 75 discordances across 8130 comparisons (allele, potential, and group levels: 5922, 1469, and 664) (supplemental Information). European samples exhibited the highest concordance (99.9% HLA-DQB1 to 97.6% HLA-DPB1; Figure 5D), while non-Europeans averaged 97.4%, with East Asian HLA-DPB1 concordance lowest at 85.7%. HLA-DPB1 comprised 41.3% of discordances. Sequencing-based retyping of 50 discordant subjects confirmed original clinical results in 48 cases. Notably, 17 (22.6%) discordances involved recently discovered HLA alleles absent from the imputation reference graph (supplemental Table 5D).

### Hereditary hemochromatosis

The UBDT_PC1 array can identify genetic variants associated with donor health, for example, 3 variants causal of hereditary haemochromatosis (HH) in the *HFE* gene: His63Asp, Ser65Cys, and Cys282Tyr.^51^ Orthogonal testing has previously validated these variants, and we observed a genotype reproducibility of ≥99.9% (Figure 2C). Among the study participants, 276 subjects had *HFE* genotypes causal of HH (supplemental Table 6). Specifically, 144 subjects were homozygous for 1 of the 3 *HFE* variants, while 132 were compound heterozygous, which causes HH if in trans. Our study confirmed that the prevalence of these HH-causing *HFE* variants differs significantly between ancestry groups and is highest in European subjects (5.8%) and absent (0%) in those of African and East Asian descent.

### Performance of UKBB_v2.2 array

Data from the 3791 samples genotyped in triplicate allowed us to assess the performance of the transfusion module when embedded into the UKBB_v2.2 array for GWAS. Genotype reproducibility between the UBDT_PC1 and UKBB_v2.2 arrays exceeded 99% for 15 762 (92.33% of 17 070 shared probes) in the unified dataset (Figure 6A). Statistical analysis revealed that 13 415 (85.1%) of the 15 762 high-reproducibility variants showed no significant difference (Pearson test, *P*≥0.001) between the MAF derived from array genotypes versus the one derived from genotypes obtained by whole genome sequencing (Figure 6B). Comparisons of the array versus clinical HEA and HPA typing results for the 3791 samples genotyped by NHSBT showed an overall concordance of 99.88% (76 829 across 76 920 comparisons) (Figure 6C, supplemental Figure 1). Of the 91 discordances, 78 were identical to those observed with the UBDT_PC1 array, with 6 and 7 additional discordances in the RH and MNS systems, respectively. Further analysis by orthogonal genotyping methods showed that all 13 were due to incorrect array results. Overall concordance between clinical and array imputed HLA types was 99.0%, with 75 discordances observed in 7435 comparisons (5671, 1471, and 218 at allele, potential, and group level, respectively). Complete overlap in HLA discordances was observed between this subset of samples and the 6679 main study samples.

**Figure 6:**
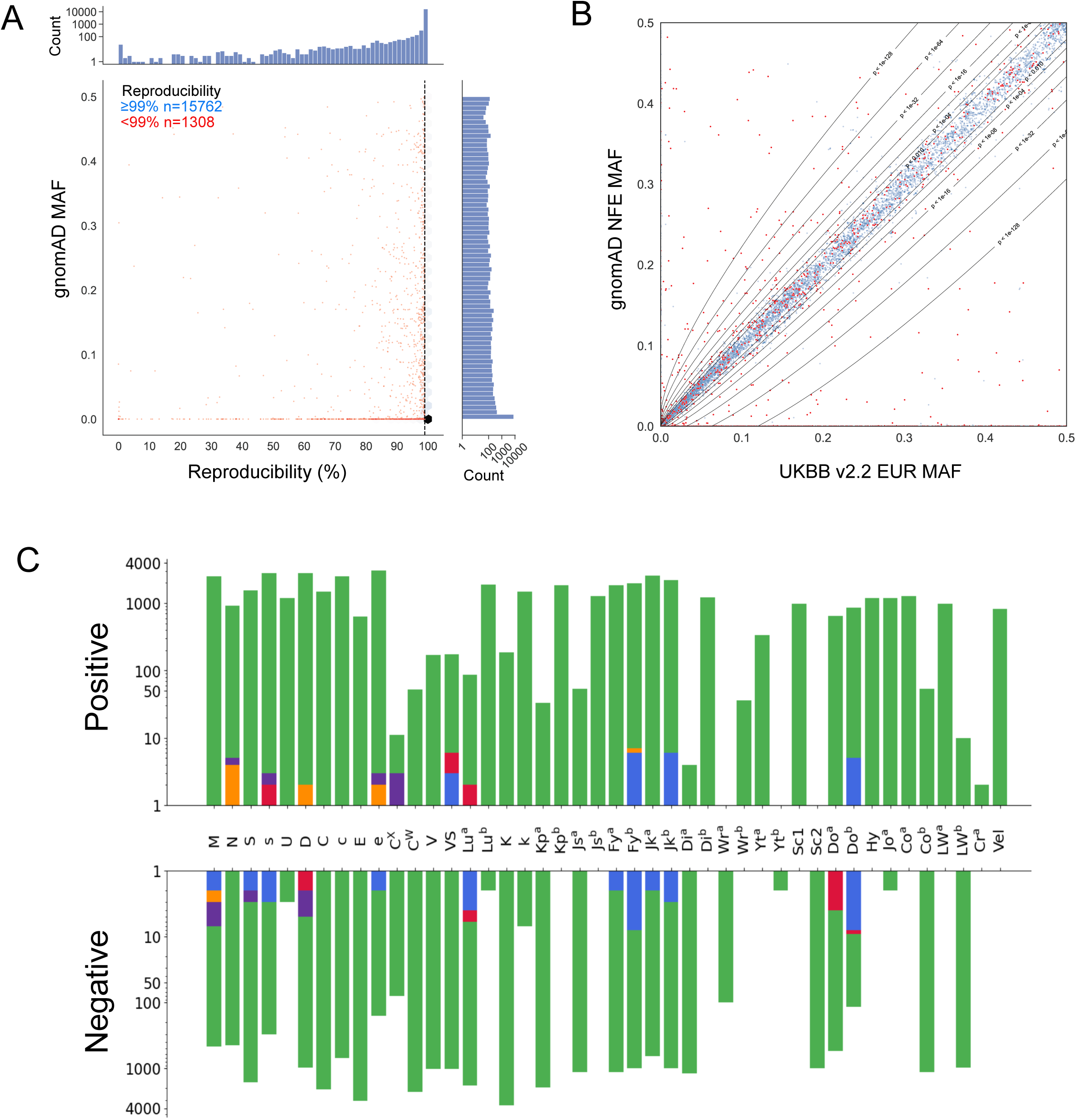
Performance of transfusion module on UKBB_v2.2 array. (**A**) Genotype reproducibility for 17 070 bi-allelic probe-variant pairs included in the UKBB_v2.2 array design. Reproducibility expressed as percentage of concordant genotype comparisons, and gnomAD minor allele frequency (MAF) for each variant are displayed on the x- and y-axes, respectively. Blue hexagons and red dots on the central scatterplot represent the density of probes with reproducibility ≥99% and individual probes with <99% concordance, respectively. Marginal histograms show probe counts on a log scale. (B) Correlation of the MAF in EUR study subjects versus (non-Finnish) EUR subjects from the gnomAD database for each probe-variant pair. Probes with ≥99% and <99% genotype reproducibility are shown in blue and red, respectively. Contour lines represent boundaries of statistical significance with corresponding P values calculated using the X-square test. (C) Concordance between clinical and array-generated Human Erythroid Antigen (HEA) types for the unified samples genotyped in triplicate (n=3791). Bar plots showing the number of comparisons (y-axis) per HEA type (x-axis) with concordant results obtained by all three test sites in green and discordant results by all test sites in blue, Sanquin only in orange, NYBC only in red, and NHSBT only in purple. Ascending and descending bars represent the number of comparisons to positive clinical or negative clinical antigen types, respectively. Bar plots show the number of comparisons on a log scale.

## Discussion

Prevention of alloimmunization through extended blood matching remains challenging in transfusion medicine due to the lack or cost of high-throughput typing methods. Using DNA from 7279 donors from 7 blood supply organizations, we demonstrate that the UBDT_PC1 transfusion-focused array effectively types 51 HEA, 8 HPA, and 6 HLA loci with high reproducibility and accuracy. When integrating the same transfusion content into the UKBB_v2.2 array designed for GWAS, performance was maintained.

To our knowledge, this is the first study to report large-scale parallel genotyping results of ancestrally diverse samples collected across multiple countries in accredited blood service laboratories. Reproducibility of test results generated on the UBDT_PC1 array and the UKBB_v2.2 array between laboratories exceeded 99% for 17 244 probes, which included a core set of antigen typing variants. Concordant antigen genotyping results across all test sites were 99.98% for HEA (n=338 372), 99.88% for HPA (n=53 270), and 99.93% for HLA (n=107 094).

Compared to available donor record HEA clinical typing results, the array-determined HEA types showed high concordance rates of 99.91% at Sanquin and 99.89% at NYBC (n=123 520 and n=123 354 HEA comparisons, respectively). Investigation of 155 discordant results supported the array-generated results in 54.2% of the cases. 37.4% were attributed to probe or algorithm performance, which can be improved by redefining probes or algorithmically. Of the remaining 8.4% miscellaneous discordances, only 2 cases were unresolved.

A lower reproducibility and higher missingness (no-type results) rates for some antigens, particularly M/N types, in African and Admixed American subjects were observed. This limitation can be resolved through algorithmic improvements that account for *GYPB* copy number when calling genotypes in the *GYPA* locus - similar to methods successfully used in RH antigen genotyping.^33^ Upon investigation, c and e antigen missingness was caused by a software issue, not array performance, and can be reduced to 2.03% and 0.23%, respectively.

A strength of this study is the population diversity, with over one-third (35.2%) of subjects being of non-European ancestry. Given the ancestral differences in blood cell antigen types, this was crucial to address the gap in previous studies focused on European populations.^45–49^ This allowed us to demonstrate detection of weak D and Del *RHD* alleles, and some complex *RHD* alleles, highlighting the array’s capability to detect D-positive donors potentially missed by serologic typing, as well as the detection of variant C antigen in subjects of African ancestry.^52,53^

The ability of the array to detect rare blood types, particularly those lacking expression of common antigens, i.e. HFA-negative types, offer the important potential to expand rare donor registries. We detected 16 uncommon or rare HFA-negative types and identified 73 new HFA-negative donors. The validation of rare antigen types remains challenging due to the limited availability of subjects with clinical typing results. However, this constraint will diminish as array testing expands to more subjects available for post-identification confirmation.

Clinical and array-imputed HLA types showed 99.1% overall concordance. European samples had the highest concordance (97.6-99.9%), while non-European samples (21.4% of total) averaged 97.4%, with East Asian HLA-DPB1 showing the lowest at 85.7%. These findings highlight the need for more diverse imputation reference data, with several ongoing studies addressing this issue using whole genome sequencing data from the UK Biobank.^36^ The capability to type class I and II HLA loci at two-field resolution alongside HEA and HPA provides a solution for delivering improved platelet transfusion care for refractory patients, offers potential for reducing rejection in kidney transplant patients requiring blood transfusions, and enriching stem cell registries with uncommon or rare HLA haplotypes, while supporting broader transfusion needs.^18,19^

Beyond antigen typing, high-density array platforms can genotype variants associated with donor health. This study identified 276 subjects with likely HH-associated genotypes, exemplifying the platform’s potential for concurrent donor health screening. The HH-genotype frequencies mirror those previously reported in the UK Biobank study, suggesting the absence of enrichment of HH carriers in NHSBT’s donor registry.^51^ The results of the array can also be used to calculate a polygenic score for ferritin using 35 independently associated variants present on the array, allowing for the incorporation of genetic effects on iron-deficiency anemia to be included in donor management.^54^

This large-scale multicentre study demonstrates that the UBDT_PC1 and UKBB_v2.2 arrays achieve exceptional accuracy and reproducibility for extended blood typing across diverse populations. While certain limitations persist, these are addressable through algorithmic improvements and expanded reference data. Validating the variants for HNA typing is part of the next development phase. The ability to simultaneously type HEA, HPA, HNA, and HLA systems, including rare antigens and donor health-related variants, while maintaining high accuracy represents a significant advance towards comprehensive matched blood product provision, especially for frequently transfused patients, like those with sickle cell disorder and thalassemia.

## Acknowledgements

The participation by blood donors as study subjects is gratefully acknowledged. Samples and data from NHSBT donors were made available through the STRIDES NIHR BioResource, which is part of the STRIDES trial. A complete list of the investigators and contributors to the STRIDES trial is provided in reference.^44^ The trial was funded by NHS Blood and Transplant and the NIHR Blood and Transplant Research Unit in Donor Health and Behaviour (NIHR203337) (formerly Donor Health and Genomics; NIHR BTRU-2014–10024) and NHS Blood & Transplant (17-01-GEN). The academic coordinating centre of STRIDES at the Department of Public Health and Primary Care at the University of Cambridge received core support from the NIHR Blood and Transplant Research Unit (NIHR203337 and NIHR BTRU-2014–10024), British Heart Foundation (RG/13/13/30194; RG/18/13/33946) and NIHR Cambridge Biomedical Research Centre (BRC-1215-20014). The NIHR BioResource receives funding from the NIHR and the NIHR Cambridge Biomedical Research Centre at the Cambridge University Hospitals NHS Foundation Trust, amongst others. Further funding for the study was received from NHSBT (grants 17-01-GEN; 20-01-GEN; G120400) for J.M. and N.S.G. and NIHR (grant G111294) for O.R. and O.S.

We thank NIHR BioResource volunteers for their participation and acknowledge the funding from NIHR. The views expressed are those of the authors and not necessarily those of the NHS, the NIHR or the Department of Health and Social Care. The support provided by the Sanquin Blood Supply Foundation and National Screening laboratory of Sanquin is much appreciated, and the funding from Sanquin to support the study is acknowledged. We also acknowledge the contribution by the Australian government’s funds for Lifeblood, which provides blood, blood products, and services to the Australian community. We also acknowledge the Finnish Red Cross Blood Service Biobank, and Drs. Satu Koskela and Jarmo Ritari for their help in providing samples from the Finnish blood donors. Research projects of J.P. are supported by the Finnish Research Council, VTR Funding from the Finnish Government, Cancer Foundation Finland, and Business Finland. Finally, we appreciate the support and resources provided by the South African National Blood Service.

## Authorship contributions

B.V., C.M.W., C.E.V.D.S., N.S.G., S.K., W.H.O., W.J.L. developed the study protocol; J.S. performed DNA sample logistics and confirmatory genotyping; A.J.G., C.J.B., L.K. performed the array genotyping; A.J.G., B.V., C.W., G.O., J.M., L.K., C.E.V.D.S. resolved HEA discordances and C.J.B., J.O., K.H., W.M.H. resolved HPA/HLA discordances; B.V., J.G., N.S.G., O.S., R.V., W.J.L. were responsible for probe design and selection and developed the IAP and were responsible for analysis; C.M.V, J.G., J.O., N.S.G., O.R., O.S., R.V., B.V. performed the bioinformatic and statistical analysis and prepared figures and tables; B.V., L.K., J.M., J.P., N.S.G., W.M.H. wrote the manuscript, which was edited by A.H., C.M.W., C.E.V.D.S., J.P., S.K., W.H.O., W.J.L.; E.A. and O.S. performed data analysis; E.D.A., K.E.S., N.K., D.R. enrolled and made available samples and data from NHSBT donors enrolled in the STRIDES NIHR BioResource.

## Disclosure of conflicts of interest

Thermo Fisher Scientific (TFS) provides research funding to the Blood transfusion Genomics Consortium and is one of its founding members. J.G. and R.V. are TFS employees and N.S.G. and W.J.L. have consultancy agreements with TFS to provide computational and scientific support for research and development. A complete list of the BGC members appears in the supplemental Information. E.D.A. holds an NIHR Senior Investigator Award.

Correspondence: Nicholas S. Gleadall, Department of Haematology, Victor Phillip Dahdaleh Heart and Lung Research Institute, University of Cambridge, Cambridge Biomedical Campus, Papworth Road, Cambridge, CB2 0BB, United Kingdom;

## Footnotes

*N.S.G and L.K. are joint first authors

‡ B.V. and W.J.L. are joint last authors

Data analysis scripts will be shared on reasonable request from the corresponding author, Nicholas S. Gleadall.

The current version of the arrays described in this manuscript are for research use only and not for use in diagnostic procedures.

---

**Supplemental Figure 1:**
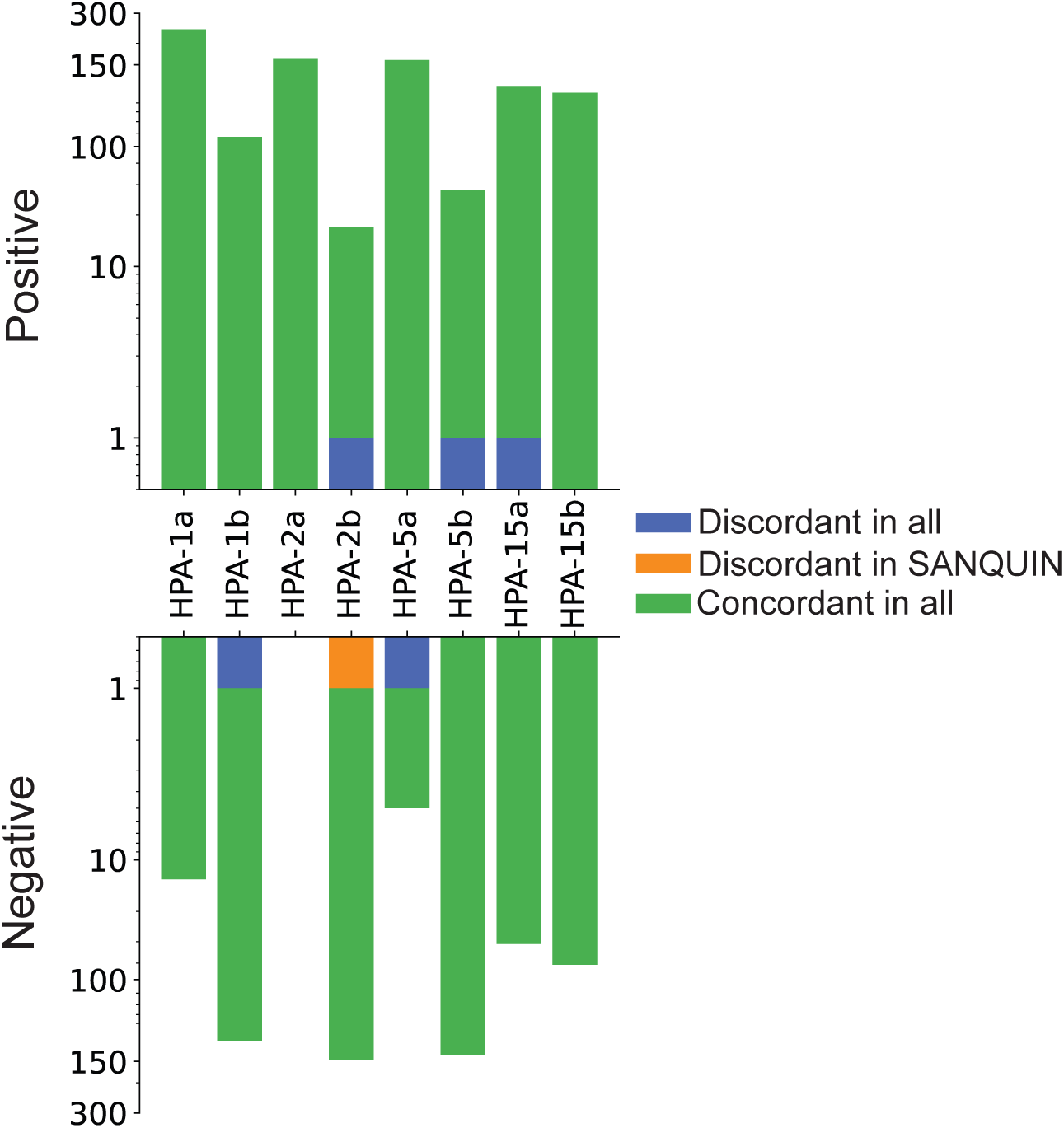
Concordance between clinical and array-generated Human Platelet Antigen (HPA) types for 6679 samples. Bar plots show the number of comparisons (y-axis) per HPA type (x-axis). Ascending and descending bars represent the number of comparisons to positive clinical or negative clinical HPA types, respectively. Bar plots show the number of comparisons on a log scale.

**Supplemental Figure 2:**
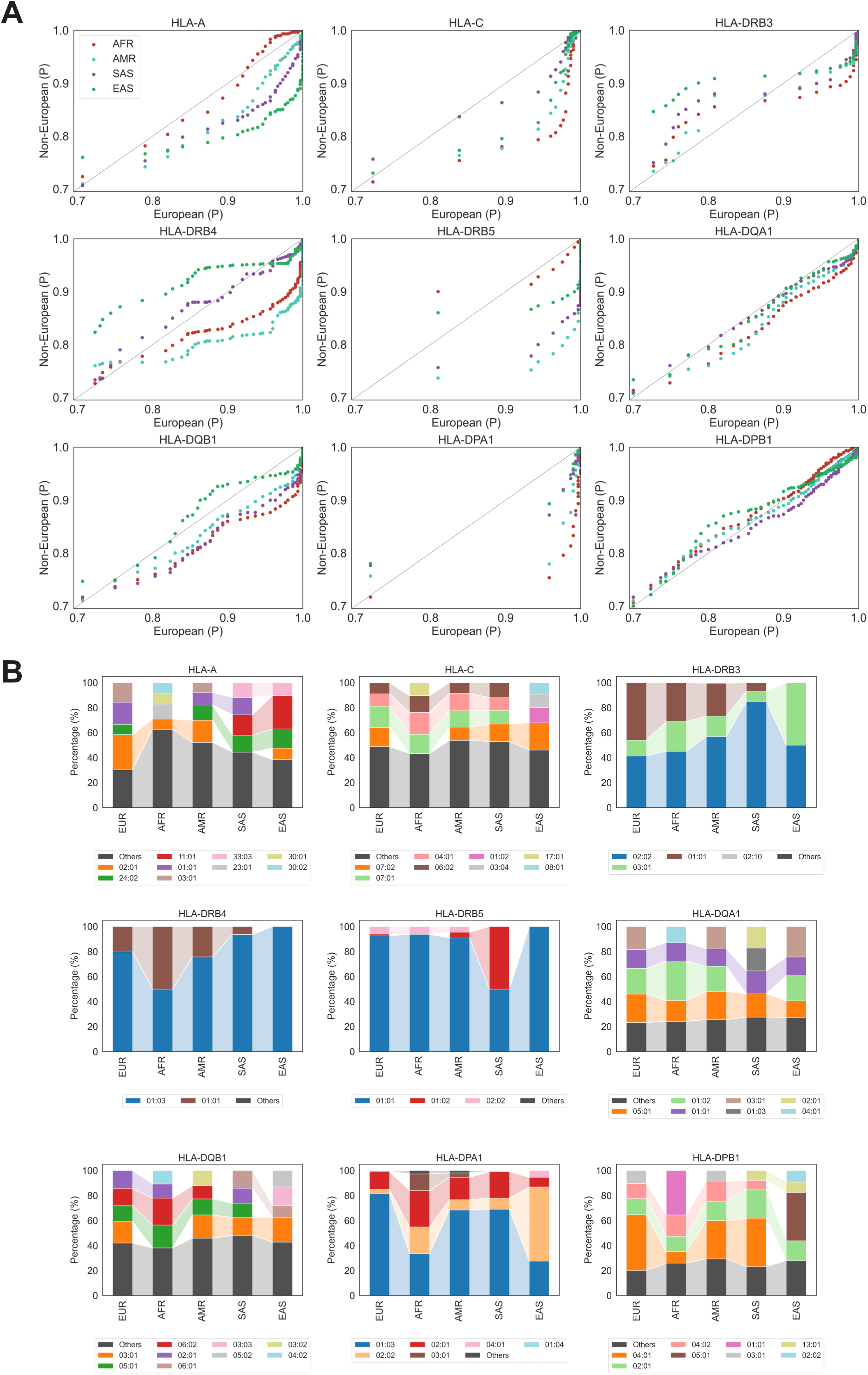
Probability distributions and diversity of UBDT_PC1 array Human Leukocyte Antigen typing data. (**A**) Quantile-Quantile (Q-Q) plots illustrating the distribution of probability scores in calling alleles for European and non-European ancestries [African (AFR), Admixed American (AMR), South Asian (SAS), and East Asian (EAS)] for the following Human Leukocyte Antigen (HLA) loci: HLA-A, HLA-C, HLA-DRB3, HLA-DRB4, HLA-DRB5, HLA-DQA1, HLA-DQB1, HLA-DPA1, HLA-DPB1, with the quantiles of probabilities (P) for European samples on the x-axis and those for non-European samples on the y-axis. (**B**) Stacked bar charts showing the frequency distribution of the top 4 alleles for the 9 HLA genes given above in (A). For the 9 HLA genes, the frequencies of the 4 top alleles are normalised to % values (y-axis) and ancestry groups are given on the x-axis. Shaded lines are drawn between bar segments representing the same allele.

## Notes

### Author Declarations

Permission for the use of DNA samples and accompanying data was obtained from the respective internal review boards (IRB) for ARCLB (Daly-08052020); CBS (2020.062-REB-20210225); FRCBS (002-2018-1); NYBC (NY-JA-190 - SOP-0210); Sanquin (EAR-250920) and SANBS (SANBS-150920). NHS research ethics committee (REC) approval was granted for the use of the samples and data from NHSBT blood donors, who were enrolled in the STRIDES NIHR BioResource Research Tissue Bank (REC-22/EE/0230; NIHR BioResource studies: NBR89 and NBR179). Samples and data from Finnish subjects were obtained from the Blood Service Biobank of the FRCBS. Permission to execute the multicentre study was approved by the IRB from Thermo Fisher Scientific. The exchange of samples and study data between Members participating in the study is regulated by the Consortium and Members' Agreement for the BGC. The data used in our study were at the individual level. However, all data were properly de-identified before their use in this study. The blood service organisations that provided the samples were responsible for anonymising the data, and these organisations held all link tables connecting samples to personally identifiable information securely. Our research team at the BGC did not collect or have access to any personally identifiable data at any point during the study.

