## Supplemental Information for "Array Genotyping of Transfusion Relevant Blood Cell Antigens in 6946 Ancestrally Diverse Subjects"

Consortium

The Blood transfusion Genomics Consortium (BGC) has 18 member organisations (members hereafter) from 14 countries. Samples and metadata used in this study were provided by 7 members from the NHSBT (UK), Sanquin (the Netherlands), NYBC (USA), ARCLB (Australia), CBS (Canada), FRCBS (Finland) and SANBS (South Africa) (supplemental Table 1).

Consent

Permission for the use of DNA samples and accompanying data was obtained from the respective internal review boards (IRB) for ARCLB (Daly-08052020); CBS (2020.062-REB-20210225); FRCBS (002-2018-1); NYBC (NY-JA-190 - SOP-0210); Sanquin (EAR-250920) and SANBS (SANBS-150920). NHS research ethics committee (REC) approval was granted for the use of the samples and data from NHSBT blood donors, who were enrolled in the STRIDES NIHR BioResource Research Tissue Bank (REC-22/EE/0230; NIHR BioResource studies: NBR89 and NBR179). Samples and data from Finnish subjects were obtained from the Blood Service Biobank of the FRCBS. Permission to execute the multicentre study was approved by the IRB from Thermo Fisher Scientific. The exchange of samples and study data between Members participating in the study is regulated by the Consortium and Members’ Agreement for the BGC.

Genotyping Laboratory Accreditation Status

The NHSBT and Sanquin laboratories are ISO 15189 accredited, and the NYBC laboratory is accredited by the American Association for the Advancement of Blood & Biotherapies and is CLIA recognised.

Sample collection and DNA quality control

Samples of EDTA anticoagulated blood were collected from non-remunerated blood donors registered with 7 blood service members of the BGC. DNA was extracted from the blood locally using accredited methods (supplemental Table 2A). DNA samples were transferred to NHSBT Cambridge for quality control (QC) and storage at -20**°**C (supplemental Table 2B). DNA quantity was measured using the Qubit Broad Range dsDNA assay kit (Thermo Fisher Scientific, Q32853). The assay results were read with the Qubit 3 Fluorometer (Thermo Fisher Scientific, Q33216). Quantified DNA samples were visually assessed for integrity by agarose gel electrophoresis using 1.0% UltraPure™ Agarose (Thermo Fisher Scientific, 16500-100). DNA was visualised with SYBR™ Safe DNA gel stain (Thermo Fisher Scientific, S33102) on a UV transilluminator. DNA samples exceeding 10 ng/µL concentration and showing no degradation upon gel electrophoresis inspection were considered to pass DNA QC and selected for further analysis.

In total, 6946 (96.6%) of the 7193 DNA samples passed DNA QC and were used for the study. For genotyping, samples were aliquoted into 96-well plates (96-Well Semi-Skirted Plates, Flat Deck, catalogue 10741725, Thermo Fisher Scientific), with each well containing 30 µl of DNA at 15 ng/µl, and each plate containing 94 study samples and 2 QC samples from the UK Blood Services Common Control Collection with identifiers UKBS0001 and UKBS0009, which were placed in wells A12 and E12 of each plate, respectively.^1^ Two identical sets of 6946 aliquots were plated for genotyping with the UBDT_PC1 array by Sanquin (Amsterdam, the Netherlands) and NYBC (Kansas City, MO, USA). Furthermore, the consent for 3938 of the 6946 samples allowed for genome-wide typing with the UKBB_v2.2 array by NHSBT (Colindale, London, UK) (Figure 1A,C). Plates were dispatched on dry ice to the accredited blood service typing laboratories and stored at -20°C until use.

Additional DNA samples

Additional sample selection (n=333) was performed independently by each center, resulting in a diverse cohort comprising both donor and patient samples. DNA from 333 subjects with rare or complex HEA or HPA phenotypes were retrieved from DNA repositories at NHSBT (n=156), Sanquin (n=90), and NYBC (n=87) and genotyped in this study to supplement the primary study sample collection described above, which lacked sufficient representation of some of the rare or complex genotypes (e.g. U−).. Due to consent limitations, these DNA samples were genotyped locally, by either NHSBT, Sanquin or NYBC, respectively (supplemental Table 2C).

Clinical typing data and sample metadata

HEA, HPA, and HLA typing results and subject metadata (e.g. data provider, self-declared gender, self-declared ancestry) were retrieved from electronic donor records and transferred via sFTP to the study database in the BGC secure research compute platform (SRCP) hosted by University of Cambridge. Hereafter, we refer to these typing results as clinical typing data. Typing for HEA was performed by a combination of haemagglutination- and DNA-based tests and HPA and HLA types were obtained using multiple DNA-based methods (supplemental Table 3A).

Array typing and quality control of genotypes

For array genotyping of the study samples, the Axiom^TM^ Propel 384 HT Workflow was followed, using the Reagents Kit 2x384HT (Thermo Fisher Scientific, catalogue 952352).^2^ 150 ng whole genome amplified target DNA was enzymatically fragmented and precipitated with isopropanol. The pelleted DNA was resuspended in a hybridisation buffer using Multidrop Combi Reagent Dispenser stations (Thermo Fisher Scientific). At Sanquin and NYBC, the hybridization-ready DNA was transferred from four 96-well microtitre plates into a single 384-well PCR plate using the ViaFlo pipetting system (Integra Biosciences) and 384 denatured DNA samples were hybridised to the UBDT_PC1 array at 48°C for 24 hours. At NHSBT, a 96-well format of the UKBB_v2.2 array was used. Following hybridisation, the array was loaded into GeneTitan™ Multi-Channel (GeneTitan hereafter) instruments (Thermo Fisher Scientific) for automated ligation, staining, washing, imaging, and genotype data generation.

Genotype quality control and antigen typing

For each DNA sample, the GeneTitan instrument generated a .CEL file containing fluorescence intensity data for individual array probes and array metadata. These files were transferred via sFTP protocol to the SRCP for analysis by the IAP, which applies multiple automated sequential modules, for genotype QC followed by antigen type calling. The following genotype QC steps were applied: (i) genotypes on a per-sample basis were QC-ed by the Axiom^TM^ Best Practices QC module; (ii) samples contaminated with >2% of DNA from another subject were identified; (iii) discordances between self-declared gender and genetically inferred sex (gender-vs-sex) were determined.^3,4^ Samples that failed any of the 3 QC steps were removed from further analysis, and the remaining QC-compliant genotypes were converted to Variant Call Format v4.2 and used for input to the bloodTyper and Applied Biosystems HLA Analysis v2.12.0RC1 algorithms to infer HEA/HPA and HLA class I and II types, respectively.^5-8^ HLA types were imputed from the genotyping data using the HLA*IMP:02 model and a multi-population reference panel of HLA haplotypes as reference.^7^ HLA typing results with a confidence score ≥0.7 were included in the study.

Genotype reproducibility

We calculated genotype reproducibility for all 20 681 probes on the UBDT_PC1 array using 6679 samples that passed genotype QC at both Sanquin and NYBC by computing the number of identical calls divided by the total number of compared genotypes. Only positions with called genotypes in both replicates were included in the analysis. The reproducibility score provides a quantitative measure of technical reproducibility, with values ranging between 0 to 1, where 1 indicates 100% reproducibility between replicates. This approach accounts for all possible genotype combinations while being robust to missing data. For the UKBB_v2.2 array analysis, genotypes from 3791 samples that passed QC in all 3 typing laboratories were examined. A consensus genotype was established by combining the 3 replicate genotypes for each probe and sample, and these consensus results were then compared to the NHSBT UKBB_v2.2 data using the same methodology as described above. To ensure a direct comparison between the UBDT_PC1 vs UKBB_v2.2 arrays the probe-variant pairs included in the analysis were restricted to those of the transfusion module.

Antigen type reproducibility

HEA and HPA typing

For each of the 51 HEA and 8 HPA types, binary confusion matrices were constructed for the 5 ancestry groups, categorising results as positive (+) or negative (−). Weak and variant HEA types were classified as positive, as is normal practice for typing of blood donors. Records with missing or no-type results from either laboratory were excluded from the reproducibility analysis. Reproducibility was computed as the sum of matching types along the diagonal of a 2x2 confusion matrix (matching positive or negative calls between laboratories) divided by the total number of compared types.

HLA typing

Four-digit (e.g. 01:01) imputed HLA types were produced for the study samples and compared first between genotyping laboratories and second to clinical types. Comparisons were grouped as follows:

Allele match: both results match for the first 2 fields (e.g. array type: *HLA-A*32:01* vs. clinical type: *HLA-A*32:01:01*).

Potential allele match: the clinical typing result is an ambiguous string of ‘potential’ alleles that contains the array-inferred typing result (e.g. array type: *HLA-A*02:01* vs. clinical type: *HLA-A*02:01*/*HLA-A*02:04*/*HLA-A*02:07*/*HLA-A*02:09*).

Group match: Both results match in the first field, but because the clinical type has not been resolved, a second field comparison cannot be made (e.g., array type: *HLA-A*24:02* vs. clinical type: *HLA-A*24*).

Mismatch: results do not agree (e.g. array type: *HLA-A*25:01* vs. clinical type *HLA-A*23:01*).

Resolution of discordances

The array-generated and clinical HEA, HPA, and HLA types were compared for all samples with valid results at Sanquin, NYBC, and NHSBT. Samples with discordant results were further analyzed by inspecting genotype call plots of specific array probes, reviewing bloodTyper software reports, evaluating clinical typing data retrieved from electronic donor record, and by conducting additional accredited molecular assays (supplemental Table 3A). ^9-13^

The NYBC laboratory investigated discordances in the MNS and RH systems. The Sanquin laboratory examined the HEA discordances for the remaining systems and investigated the HPA discordances. Where DNA was available, samples were also analyzed using an NGS panel test targeting 53 genes relevant to HEA typing. Discordances were categorized as array correct, array incorrect, algorithmic, and miscellaneous. The NHSBT resolved the discordances between array-generated and clinical HLA typing results.

Additional contributors to the Blood transfusion Genomics Consortium

Ai Leen Ang^1,2^, William J. Astle^3^, John Boucher^4,5^, John R. Bradley^4,6,7,8^, Candice Davison^9^, Tanya N. Glatt^10^, Andreas Greinacher^11^, Tammy Ison^12^, Hartirathpal Kaur Juspal Singh^2^, Shireen Jyawook^13^, Aliyye Karasu^14^, Mary Kasanicki^8^, Marco Koppelman^15^, Pawinee Kupatawintu^16^, Jennifer Laird^17^, Jacinta Lee^4^, Stefan Mayer^18^, Ana-Maria Moreno^19^, Nigel Ovington^4,5^, Brad Pfaltzgraff^20^, Lydia Quaye^14^, Luisa Ronzoni^21^, Rob Schleifer^13^, Melissa Schreiner^13^, Kathleen Selling^11^, Hannah Stark^4,5^, Sara Trompeter^22,23^, Luca Vallenti^21^, Sumathi Venkatapathy^13^, Phandee Watanaboonyongcharoen^24^

^1^Department of Hematology, Singapore General Hospital, Singapore, Singapore, ^2^Blood Services Group, Health Sciences Authority, Singapore, Singapore, ^3^MRC Biostatistics Unit, University of Cambridge, Cambridge, United Kingdom, ^4^NIHR BioResource, Cambridge, United Kingdom, ^5^Department of Haematology, Victor Phillip Dahdaleh Heart and Lung Research Institute, University of Cambridge, Cambridge, United Kingdom, ^6^NIHR Cambridge Biomedical Research Centre, Cambridge, United Kingdom, ^7^Addenbrookes Hospital, Cambridge University Hospitals NHS Foundation Trust, Cambridge, United Kingdom, ^8^Cambridge University Hospitals, Cambridge, United Kingdom, ^9^Pathology and Clinical Governance, Australian Red Cross LifeBlood, Brisbane, Australia, ^10^Immunohaematology Reference Laboratories, South African National Blood Service, Johannesburg, South Africa, ^11^Institute for Immunology and Transfusion Medicine, University Medicine Greifswald, Greifswald, Germany, ^12^Canadian Blood Services National Reference Laboratory, Canadian Blood Services, Brampton, Ontario, Canada, ^13^Microarray Solutions, Thermo Fisher Scientific, Santa Clara, CA, United States, ^14^Histocompatibility and Immunogenetics laboratory, National Health Service Blood and Transplant, London, United Kingdom, ^15^National Screening laboratory of Sanquin, Sanquin Blood Supply Foundation, Sanquin, Amsterdam, the Netherlands, ^16^National Blood Center, Thai Red Cross Society, Bangkok, Thailand, ^17^Red Cell Immunohaematology, Scottish National Blood Transfusion Service, Glasgow, United Kingdom, ^18^Department of Molecular Diagnostics and Cytometry, Blood Transfusion Service Zurich, Swiss Red Cross, Zurich, Switzerland, ^19^Pathology and Clinical Governance, Australian Red Cross Lifeblood, Brisbane, Australia, ^20^National Center for Blood Group Genomics, New York Blood Center Enterprise, Kansas City, MO, United States, ^21^Department of Transfusion Medicine and Hematology, Policlinico of Milan, Milan, Italy, ^22^University College London Hospitals NHS Foundation Trust, London, United Kingdom, ^23^National Health Service Blood and Transplant, London, United Kingdom, ^24^Department of Laboratory Medicine, Chulalonkorn University Transfusion Medicine Unit, King Chulalongkorn Memorial Hospital, Bangkok, Thailand
